# ELECTROCLINICAL CHARACTERISTICS, GENETIC FINDINGS AND COMPLEX EPILEPSY SURGERY OUTCOMES IN CHILDREN WITH LOW-GRADE EPILEPSY-ASSOCIATED TUMORS – A COMPREHENSIVE VIEW

**DOI:** 10.1101/2025.04.28.25324638

**Authors:** Gonzalo Alonso Rivera Ramos, Barbora Straka, Petr Jezdik, Alice Maulisova, Katerina Bukacova, Martin Kudr, Alena Jahodova, Anezka Belohlavkova, Martin Kyncl, Zuzana Holubova, Radek Janča, Miroslav Koblizek, Josef Zamecnik, Lenka Krskova, Martina Strnadova, Michal Zapotocky, Michal Tichy, Vladimír Beneš, Petr Liby, Pavel Krsek

**Author notes:** **Corresponding author’s contact information** Pavel Krsek, MD, Department of Paediatric Neurology, Second Faculty of Medicine and Motol University Hospital, Prague, Czech Republic. **Author contribution statement** Gonzalo Alonso Ramos Rivera and Pavel Kršek were involved in conceptualization, investigation, data curation, and writing - original draft preparation. Barbora Straka and Radek Janča were involved in writing, review & editing. Martin Kudr, Petr Jezdik, Alena Jahodova, Anezka Belohlavkova, Katerina Bukacova, Alice Maulisova, Miroslav Koblizek, Josef Zamecnik, Zuzana Holubova, Martin Kyncl, Petr Liby, Michal Tichy, Vladimír Beneš, Lenka Krskova, Martina Strnadova, Michal Zapotocky were involved in resources & final text editing. **Study approval statement** The study was approved by the institutional Ethical committee of Motol University Hospital (2022/06/15 - EK - 602.24/22). **Patient consent statement** Patients and/or their legal representatives gave consent with the study. **Ethical publication statement** We confirm that we have read the Journal’s position on issues involved in ethical publication and affirm that this report is consistent with those guidelines.

## Abstract

**Objective:** To analyze postsurgical outcomes in relation to epilepsy characteristics and genetic etiology in pediatric patients with isolated low-grade epilepsy associated tumors (LEAT) and LEAT plus focal cortical dysplasia type IIIb (FCD IIIb) who underwent epilepsy surgery.

**Methods:** Patients younger than 19 years at the time of epilepsy surgery, with isolated LEAT or LEAT plus FCD IIIb and a minimum follow-up of 2 years were included. Clinical data, neuroimaging, EEG, neuropsychological findings, surgical variables, histopathological and molecular-genetic findings were evaluated. Surgical outcomes were assessed in four domains: seizures, antiseizure medication (ASM) use, cognitive performance changes and complications, including predictor analysis.

**Results:** Seventy-three children fulfilled the inclusion criteria, with 53 (72.6%) having drug-resistant epilepsy. LEAT plus FCD IIIb were more frequent than isolated LEAT (44/73, 60.3% vs. 29/73, 39.7%) and gangliogliomas were the most common tumor type (43/73, 58.9%), followed by dysembryoplastic neuroepithelial tumor (19/73, 26.0 %). Genetic testing was more frequently positive in isolated LEAT (20/28) than LEAT plus FCD IIIb (18/43, p = 0.02). At the end of follow-up (median 5.7 years), 66 patients (90.4%) were seizure-free, and 57 (78.1%) had discontinued ASM. Younger age at seizure onset, longer epilepsy duration, and a higher number of ASM were correlated to lower pre- and postoperative IQ. An IQ gain of > 10 pts. postoperatively was present in 8 patients (15.1%). Two patients (2.7%) had an unexpected permanent deficit, while 10 (13.7%) had minor temporary deficit. No additional predictive outcome factors were identified.

**Significance:** Epilepsy surgery yields high chances of freedom from seizures and ASM discontinuation. Early surgical intervention in terms of shorter duration of epilepsy and fewer used ASM can be associated to a higher pre- and postoperative IQ. Molecular-genetic differences between isolated LEAT and LEAT plus FCD IIIb suggest distinct neoplastic and dysplastic entities, respectively.

**Key points:**

1. A majority of patients with LEAT achieved freedom from seizures and ASM post-surgically.
2. Children with younger age at seizure onset, longer duration of epilepsy and a higher number of ASM used before surgery had lower pre- and postoperative IQ.
3. Genetic cause was detected more often in patients with isolated LEAT vs. LEAT + FCD IIIb implying their neoplastic respectively dysplastic origin.

## 1. INTRODUCTION

Tumors are one of the most common causes of seizures and epilepsy. Among them, the so-called “low-grade epilepsy associated tumors” (LEAT) represent the most important group. LEAT are, from the histopathological point of view, low-grade glioneuronal tumors with a rare tendency towards malignant progression. They comprise mostly gangliogliomas (GG) and dysembryoplastic neuroepithelial tumors (DNET). However, LEAT spectrum is broad and includes papillary glio-neuronal tumor, angiocentric gliomas, isomorphic astrocytoma, polymorphous low-grade neuroepithelial tumor of the young (PLNTY), and others. Focal cortical dysplasia (FCD) is frequently associated with LEAT, defined as FCD type IIIb^1^. It is important to note that the term “LEAT” refers to the association of the respective tumor with the diagnosis of drug-resistant epilepsy (DRE) as the dominant clinical manifestation, and not to a specific histopathological diagnosis.

The importance of epilepsy surgery in LEAT was clearly demonstrated by Lamberink *et al*. ^2^ on a large multicenter cohort of more than 9100 patients, with almost 1500 patients having LEAT. The authors found that among all histopathological diagnoses, patients with LEAT had the best outcome of epilepsy surgery – 80.3% of patients were assessed as Engel I after 1 year, 77.5% after 2 years and 75.9% after 5 years. Nevertheless, despite the high incidence of LEAT in epilepsy surgery cohorts, electroclinical characteristics and their relationship to outcomes other than seizure-related have not been deeply investigated, mainly in children. There exists a wide gap between a large body of knowledge on the molecular-genetic mechanisms of tumor formation and its relation to patients’ prognoses, reflected in the most recent WHO classification of brain tumours 5^th^ edition, and very limited evidence on the pathogenesis of FCD type III, i.e. FCD associated with another lesion^3^. Multiple recently published studies have evaluated the effect of genetic background on surgical outcomes in epilepsy surgery cohorts^4–6^; however, these studies focused on patients with malformations of cortical development and largely excluded patients with LEAT.

Our study presents a complex view on outcome domains after epilepsy surgery including seizures, postoperative use of ASM, changes in cognition and surgical complications. We analyzed a wide spectrum of possible predictive factors in a large unicentric pediatric surgical cohort, including genetic findings in LEAT tissue; to our knowledge, this is the first study to assess the role of genetic etiology on epilepsy surgery outcomes in patients with LEAT. We also compared electroclinical characteristics and outcomes of patients with isolated LEAT and LEAT plus FCD IIIb.

## II. MATERIALS AND METHODS

### 1. Patient selection

Based on electronic database of Motol Epilepsy Center, we selected pediatric patients who underwent epilepsy surgery between January 2002 and December 2020, and fulfilled the following selection criteria: (1) age under 19 years at surgery time, (2) confirmed histopathological finding of LEAT isolated or with FCD IIIb., (3) a minimal postsurgical follow-up of 2 years.

### 2. Preoperative evaluation, surgery approach

A detailed personal history was taken, including family history of seizures and brain tumors, age at seizure onset, seizure type, frequency of seizures, presence of infantile spasms and status epilepticus, number of ASM before surgery and, consequentially, the diagnosis of drug-resistant epilepsy^7^.

In the majority of patients, we analyzed pre-surgical IQ/DQ. A neuropsychological examination was conducted before surgery using the latest available national versions of age- and cognitively appropriate assessments: the Wechsler Adult Intelligence Scale, 3rd edition (WAIS-III)^8^, the Wechsler Intelligence Scale for Children, 3rd edition (WISC-III)^9^ for IQ evaluation, and the Bayley Scales of Infant Development, 2nd edition (BSID-II)^10^ for developmental quotient assessment. Stanford-Binet Intelligence Scale in 4th revision^11^ was selected for IQ testing in children with lower cognitive skills (expected IQ < 60), despite its limited reproducibility and reliability of IQ testing in this group of patients.

All patients underwent brain MRI (1,5 or 3T) in a dedicated epilepsy protocol, which is continuously improved in our center based on the latest findings. MR examinations for the vast majority of patients were in accordance with the recommendations of the ILAE for the use of MRI in patients with epilepsy^12^. In selected cases, MR spectroscopy, fMRI, WADA-test, DTI, 18FDG-PET, and/or SISCOM were performed, with the aim to ascertain tumor localization, its relationship to eloquent cortex, lesion characteristics and/or lesion margins. Based on the data, we determined lesion size (sublobar, lobar, multilobar), its localization (temporal, extratemporal, temporal plus), relationship to eloquent cortex, and evolution of lesion size if sequential neuroimaging was performed.

All but two patients underwent scalp long-term video EEG; however, a recording of an epileptic seizure during video EEG was not a prerequisite for resective epilepsy surgery in patients with a convincing localization of a structural lesion on MRI that correlated with the reported seizure semiology. As a standard practice in our center, intraoperative ECoG was performed in all but two patients. Patients with supposed epileptogenic zone with unclear margins and/or in proximity to the eloquent cortex, underwent long-term invasive stereo-EEG study. Most patients with a close relationship between the epileptogenic zone and eloquent cortical areas or significant white matter tracts were operated on using a combination of ECoG and intraoperative motor function mapping^13^. In rare cases, awake craniotomy was used.

Epilepsy surgery variables included age at surgery, duration of epilepsy (elapsed time from the first seizure until surgery), use of ECoG and its findings, use of long-term invasive SEEG and intraoperative monitoring (IOM), and completeness of resection according to ECoG and MRI.

### 3. Postoperative follow-up

We evaluated surgical outcomes in terms of (1) post-surgical seizure frequency using ILAE scale, (2) withdrawal of ASM, (3) cognitive outcomes (shift in full-scale IQ postoperatively) and (4) surgical complications, comprising minor (non-life-threatening and/or lasting less than 3 months) and major (life-threatening, need of surgical intervention and/or duration of more than 3 months)^14^. The assessment of seizure outcome, postoperative use of ASM and surgical complications was performed at the last recorded medical visit or by phone. Post-surgical neuropsychological assessment was performed routinely at least one year after surgery.

### 4. Histopathological and molecular-genetic analyses

Brain tissue samples obtained in epilepsy surgery were processed in accordance with published recommendations by the ILAE^15,16^. Brain tumors were classified according to the World Health Organization classification scheme for Central Nervous System Tumors (5th edition) for low-grade epilepsy-associated neuroepithelial tumors^17^. DNA was extracted from the formalin-fixed paraffin-embedded (FFPE) samples using QIAamp DNA FFPE kit (Qiagen) and from fresh-frozen samples by Trizol DNA isolation kit (Invitrogen) as per manufacturers’ protocols. Methods of molecular-genetic analyses comprised Sanger sequencing for detection of V600E pathogenic variant in *BRAF*, real-time PCR and Sanger sequencing for detection of *KIAA1549::BRAF* and internal tandem duplication of *FGFR1* and next-generation sequencing methods utilizing FusionPlex kits (Archer Dx., Boulder, USA) and Archer Dx. software.

### 5. Study design and statistical analysis

We analyzed the influence of the above-mentioned variables (demographic and clinical data, neuroimaging and neuropsychological findings, surgical variables, histopathological findings) on the different outcomes, comparing (1) patients completely seizure-free after surgery (ILAE 1) vs cases with all other seizures outcomes, (2) ASM-free vs. non-ASM-free patients, (3) patients with an increase in full-scale IQ of > 10 points vs. ≤ 10 points, and (4) patients with no / minor vs. major surgical complications, with the intention to find predictive factors of these outcomes. Similarly, we looked for predictive factors associated with pre- and postoperative IQ. Finally, we compared patients with isolated LEAT vs. LEAT+FCD IIIb.

We conducted univariate analyses to evaluate the relationship between various predictors and each outcome of interest. Continuous variables were assessed using a parametric approach with analysis of variance (ANOVA) tests to detect significant differences among groups. For categorical variables, Fisher’s exact test was employed to account for smaller sample sizes in certain categories. Although generalized linear models were initially explored to assess multivariable relationships across predictors, only univariate results are presented due to the focus on individual predictor-outcome associations. A significance level of p < 0.05 was applied throughout the analyses. To address the large number of tested predictors, p-values were adjusted using a False Discovery Rate (FDR) correction, and all p-values reported in the Results section are presented after FDR adjustment. Furthermore, for some predictors, regression analyses were conducted to gain deeper insights into their relationship with the outcomes of interest.

## III. RESULTS

### 1. Demographic and clinical data

Seventy-three subjects fulfilled the above-defined inclusion criteria, 37 boys (50.7%) and 36 girls (49.3%). Four patients (5.5%) had a positive familiar history of brain tumors, and 4 patients (5.5%) had a first- or second-degree relative with epilepsy. The median age at seizure onset was 6.0 years (range 0.1 – 16.8 years). Seizures were classified as focal in 93.2% (68 patients), including 16.4% having FBTCS (12 patients). Epileptic spasms occurred in 3 children (4.1%), and status epilepticus also in 3 patients (4.1%). Twenty-seven patients (37%) had daily seizures, 25 (34.2%) weekly and 21 (28.8%) monthly or less frequent seizures. The median number of preoperatively used ASM was 2 (range 1 – 7) and the number of ASM used at the time of surgery 2 (range 1 – 3). Fifty-three patients fulfilled the criteria for DRE (72.6%). Table 1 shows demographic and clinical data.

**Table 1.** Demographic and clinical data

|  |  |
| --- | --- |
| **Total number of patients** | 73 |
| Male | 37 (50.7%) |
| Female | 36 (49.3%) |
| **Median age at seizure onset (range)** | 6.0 years (0.1 – 16.8) |
| **Type of seizures** |  |
| Focal | 68 (93.2%) |
| FBTCS | 12 (16.4%) |
| Generalized | 5 (6.8%) |
| **Frequency of seizures** |  |
| Daily | 27 (37.0%) |
| Weekly | 25 (34.2%) |
| Monthly or less | 21 (28.8%) |
| **Patients with drug-resistant epilepsy** | 53 (72.6%) |

### 2. Preoperative evaluation

#### 2.1. Neuropsychological data

Sixty-two patients had neuropsychological evaluation before surgery and in 53 both pre- and postoperative testing were performed. Most of the remaining patients did not undergo neuropsychological tests for language reasons (non-Czech-speaking patients). Median IQ before surgery was 98 (range 44 – 124); four patients (6.4%) achieved pre-surgical FS-IQ < 70. Linear regression analyses confirmed that younger age at seizure onset (β = 0.99, p = 0.022, R^2^ = 0.084), longer duration of epilepsy (β = –1.72, p = 0.002, R^2^ = 0.16), and a higher total number of ASM used before surgery (β = –3.79, p = 0.004, R^2^ = 0.133) each served as significant predictors of lower preoperative FS-IQ (Figures 1A-C). WADA test was performed in 11.0% (8 patients).

**Figure 1.**
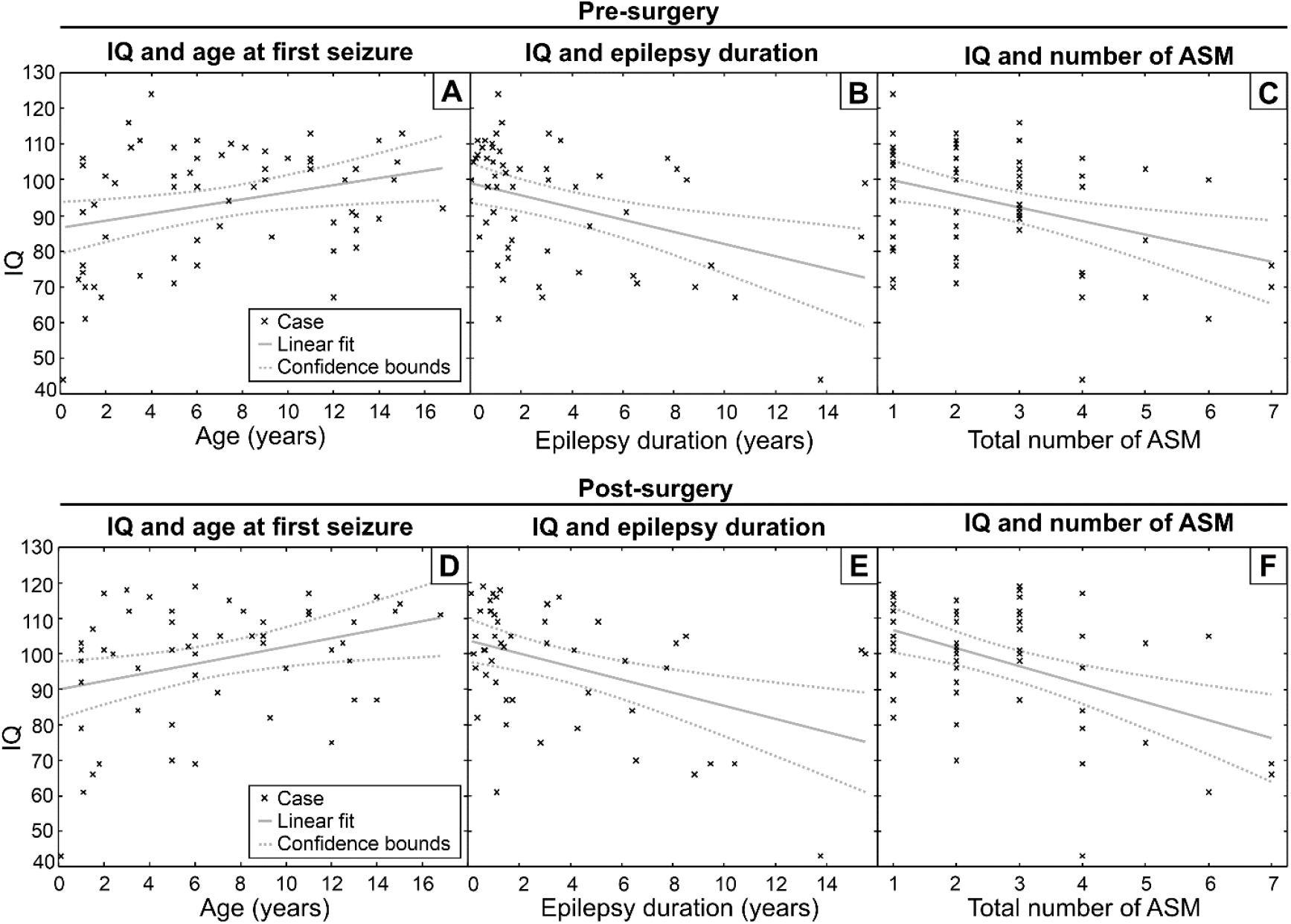
Correlation between age at seizure onset, epilepsy duration, total ASM number before surgery and pre/postsurgical FS-IQ. (A) Data for age at seizure onset vs. presurgical FS-IQ (adjusted R^2^ = 0.069, p value 0.022). (B) Data for duration of epilepsy vs. presurgical FS-IQ (adjusted R^2^ = 0.145, p value 0.002). (C) Data for ASM number vs. presurgical FS-IQ (adjusted R^2^ = 0.119, p value 0.004). (D) Data for age at seizure onset vs. postsurgical FS-IQ (adjusted R^2^ = 0.09, p value 0.016). (E) Data for duration of epilepsy vs. postsurgical FS-IQ (adjusted R^2^ = 0.173, p value 0.002). (F) Data for ASM number vs. postsurgical FS-IQ (adjusted R^2^ = 0.204, p value < 0.001).

#### 2.2. Neuroimaging data

Lesions were localized predominantly in the temporal lobe (42 patients, 57.5%). According to their size, they were classified as sublobar in 53 patients (72.6%), lobar in 10 (13.7%) and multilobar in 10 (13.7%). Of 43 patients who had sequential MRI, 27.9% (12 children) showed growth of the tumor. Additional neuroimaging methods were used as follows: MR spectroscopy in 24.7% (18 patients), fMRI in 61.6% (45 patients), DTI with the reconstruction of important white matter tracts incorporated in neuronavigation in 68.5% (50 patients), 18FDG-PET was performed in in 79.5% (58 patients), and SISCOM in 8.2% (6 patients).

#### 2.3. Scalp EEG data

Scalp EEG was performed in all patients and long-term video EEG in 71 (97.2%). Interictal epileptiform activity was regional in 56.2% of cases, multiregional in 26.0%, hemispheral in 4.1% and generalized in 2.7%. Ictal EEG showed regional onset of seizure(s) in 57.5%, multiregional (i.e. widespread ictal pattern within one hemisphere or with a fast contralateral spread) in 20.5% and generalized in 2.7%. Concordant findings of both regional interictal and ictal EEG were found in 30 patients (41.1%).

### 3. Surgical variables

The median age at epilepsy surgery was 10.1 years (2.0 – 18.9) and median duration of epilepsy 1.9 years (0.03 – 15.7). Intraoperative ECoG was performed in 71 (97.3%) including postresective ECoG in 29 cases (39.7%). In 10 cases (10/71, 14.1%) the resection was modified based on ECoG findings, see example cases in Figures 2 and 3. Long-term invasive SEEG was performed in 4 cases (5.5%). Forty-seven patients (64.6%) had a lesion in proximity of eloquent cortex and in 21 patients (28.8%) continuous intraoperative monitoring of motor functions (IOM) was performed. Awake craniotomy was done in one patient (1.4%). According to postoperative MRI, resection was considered as complete in 60 patients (83.3%). Seven patients (9.6%) were reoperated because of oncological indication (growth of a residuum). Four of them have isolated LEAT and 3 FCD IIIb.

**Figure 2.**
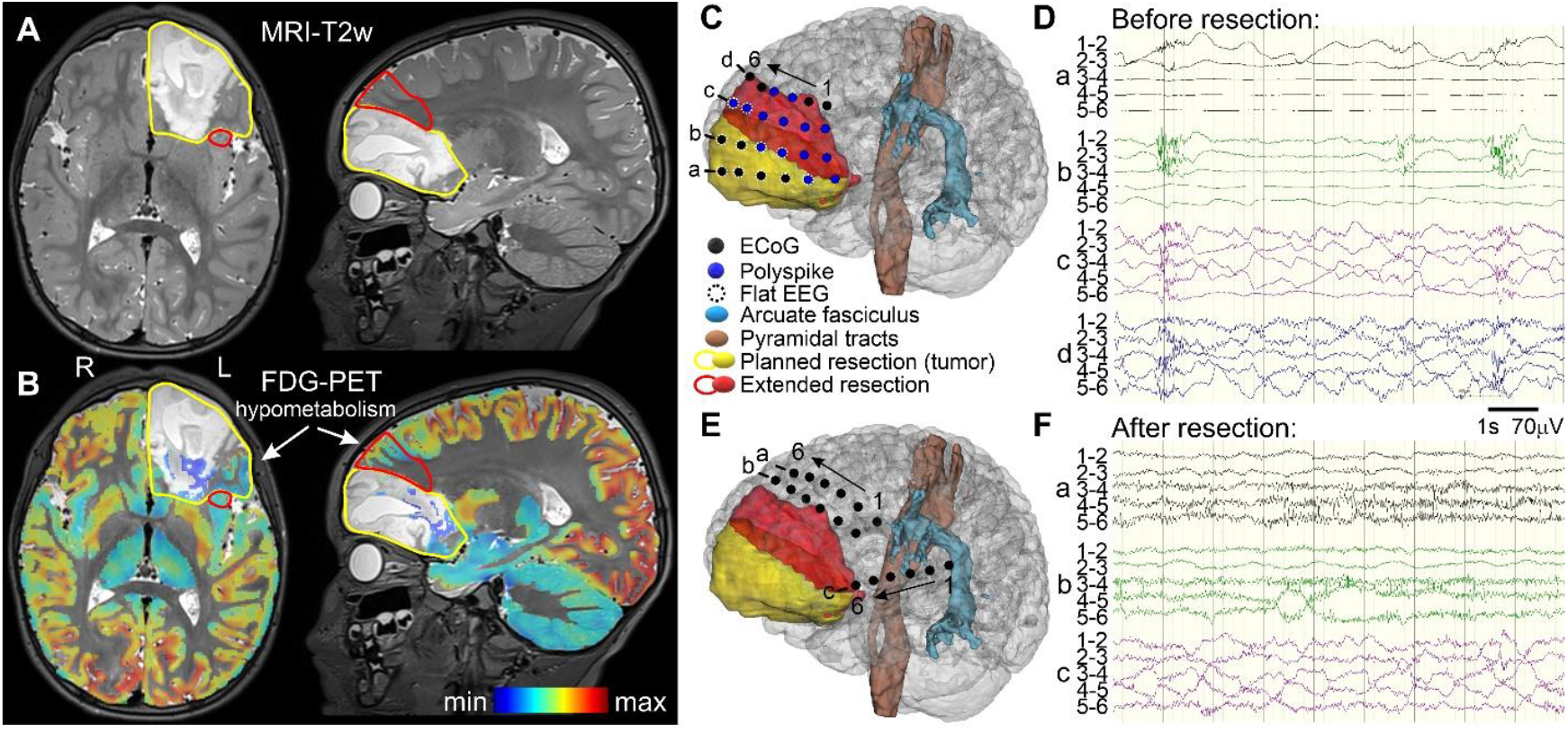
Illustrative case report of a patient with isolated LEAT. Preschooler with DRE and DNET. (A) MRI (T2w images) depicting a large tumor in the left-frontal lobe. Yellow contour marks initially planned resection, red contours marks extension of resection based on intraoperative ECoG finding. (B) FDG-PET registered with MRI was post-processed by partial volume effect correction to hypometabolism highlight^36^. (C) Three-dimensional patient visualization by 3D Slicer showing planned (yellow) and extended (red) resection. Dots represent ECoG strips placement before resection. (D) ECoG recording shows flat curve over the tumor and polyspikes generated by the hypometabolic cortex - the finding leading to the resection extension. (E) 3D visualization of ECoG strips placement after resection, which confirmed disappearance of the ECoG interictal epileptiform activity (F). The patient is postsurgically seizure-free, ASM-free, with no neurological deficit and the normal cognitive profile.

**Figure 3.**
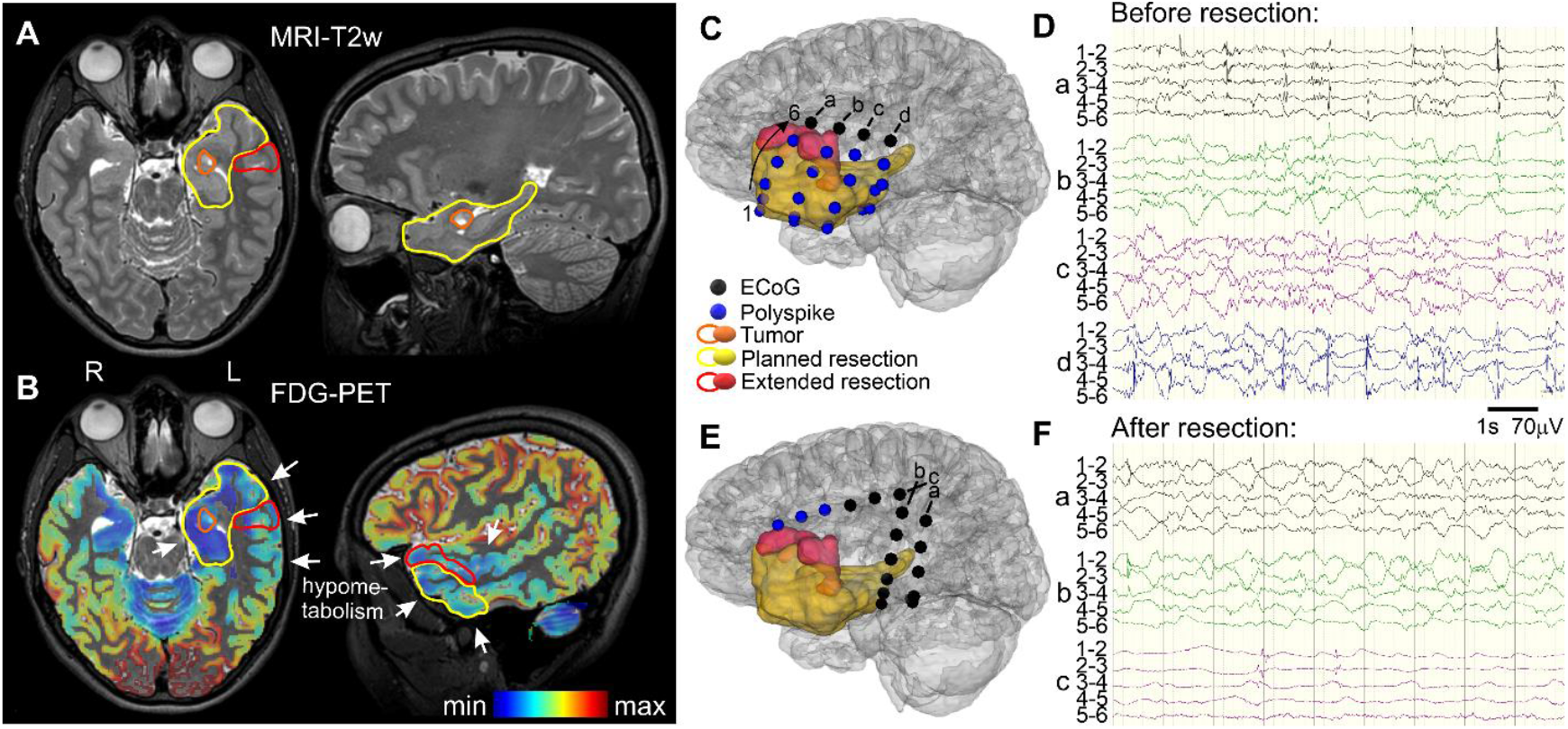
Illustrative case report of a patient with LEAT plus FCD. Adolescent with DRE and GG plus associated FCD IIIb. (A) MRI (T2w images) depicting a small tumor in left amygdala (orange contour) visible on MRI. Yellow contour marks initially planned resection that took into account suspected dysplastic changes in the surrounding mesial temporal structures, including the anterior part of the hippocampus. Red contours marks extension of resection based on intraoperative ECoG finding. (B) FDG-PET registered with MRI was post-processed by partial volume effect correction to hypometabolism highlight^36^ Hypometabolic regions well correlated with the suspected dysplastic changes on MRI. (C) Three-dimensional patient visualization by 3D Slicer: planned (yellow) and extended (red) resection. Dots represent ECoG strips placement before resection. (D) ECoG recording showed polyspikes over the hypometabolic cortex leading to the resection extension. (E) 3D visualization of ECoG strips placement after the resection, with a marked reduction of the ECoG epileptiform activity (F). The patient is postsurgically seizure-free, ASM-free, with no neurological deficit and a normal cognitive profile, studying a university.

### 4. Histopathological and molecular-genetic findings

Isolated LEAT were present in 29 patients (39.7%) and FCD IIIb in 44 (60.3%). The most frequent tumor type was GG (43 patients, 58.9%), followed by DNET (19 patients, 26.0%). Eleven patients (15.1%) had other types of tumors (Table 2). Comparing isolated LEAT and LEAT associated with FCD IIIb, significant differences were found in lesion localization – tumors associated with FCD IIIb were localized more frequently in the temporal lobe (31.0%) and isolated LEAT in extratemporal regions (69.0%, p value < 0.001); and tumor type – GG was the dominant tumor type in tumors associated with FCD IIIb (84.1%), while DNET was the most frequent type in isolated LEAT (48.3%, p value < 0.001). Positive results of genetic testing were more frequent in the cohort of isolated LEAT (positive n=20, negative n=8) compared to the cohort of LEAT with FCD IIIb (positive n=18, negative n=25; p = 0.02). Table 2 contains data on histopathological and molecular-genetic findings, detailed in Supplementary table 1.

**Table 2.**
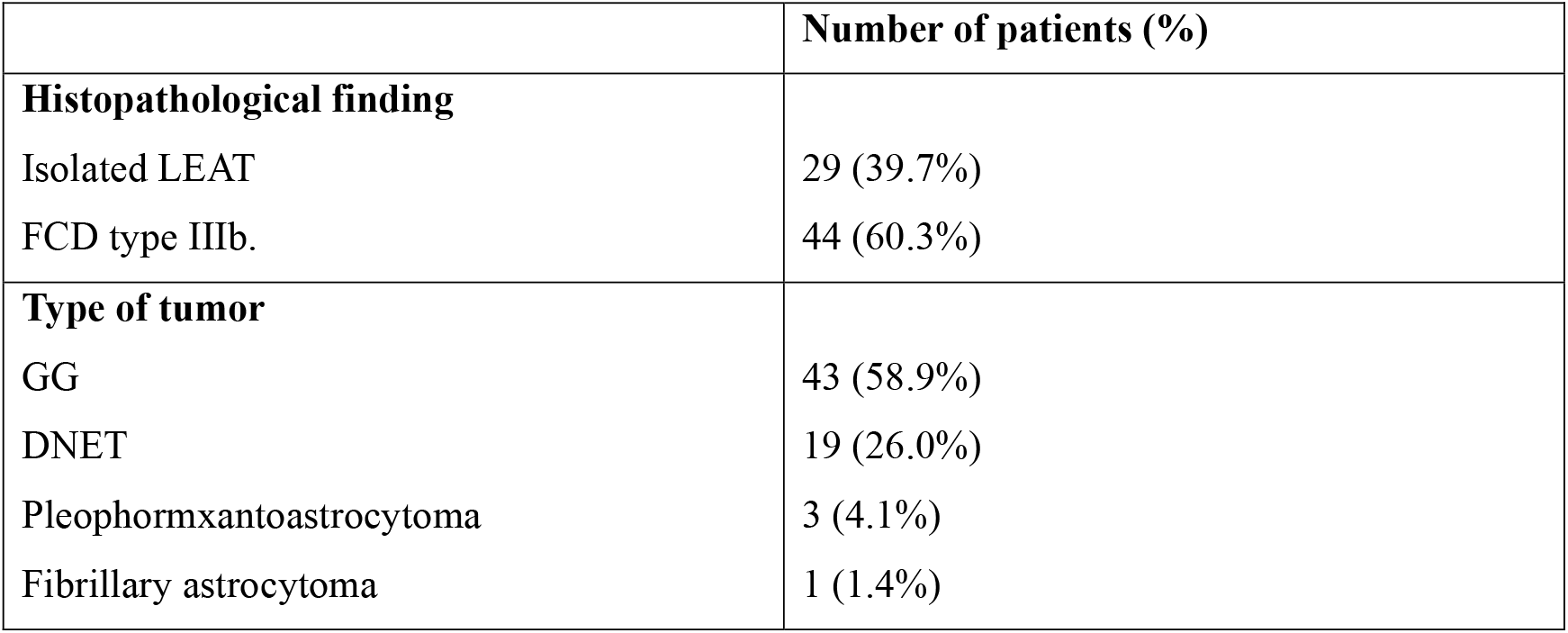
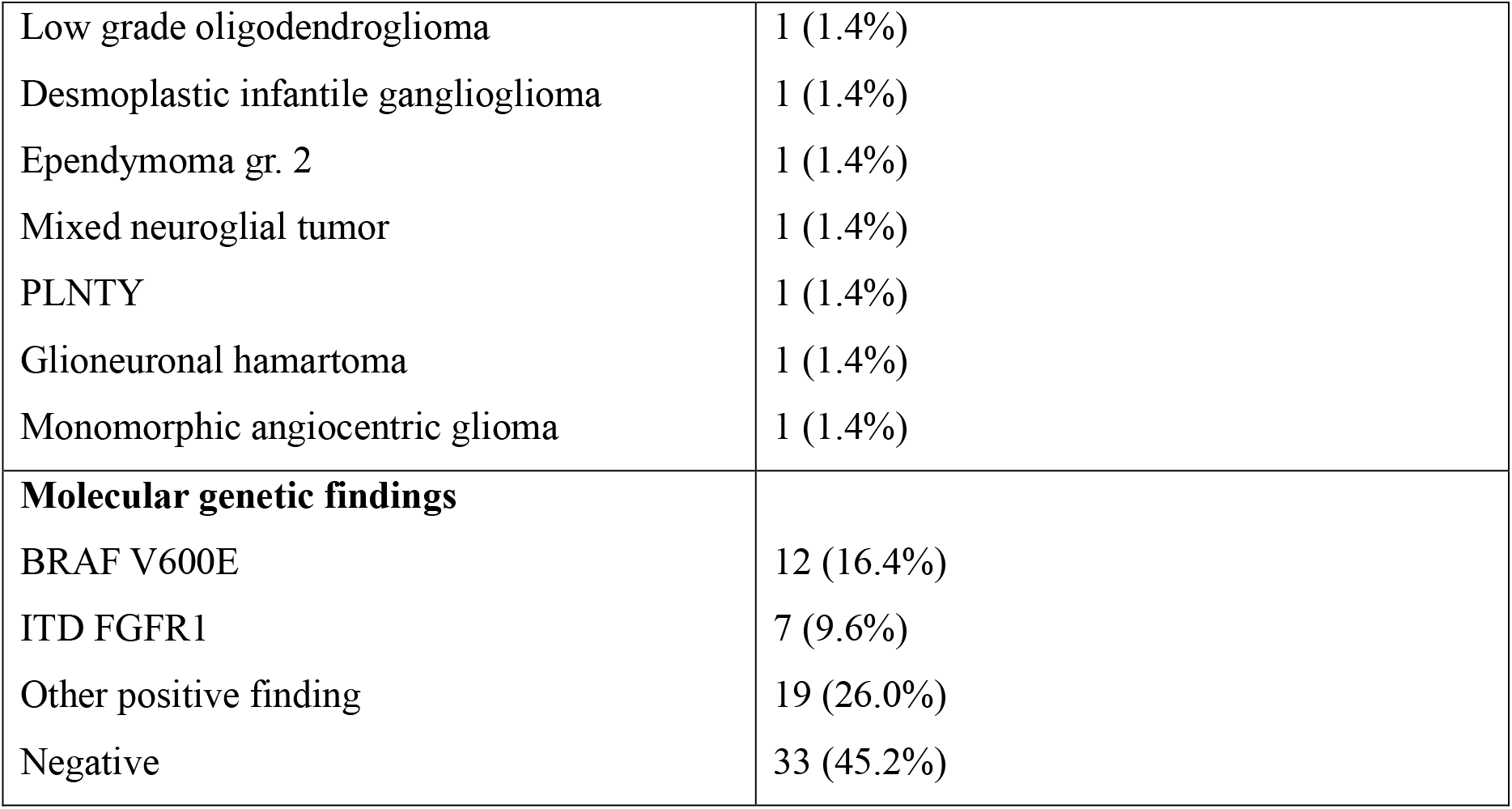
Histopathological and molecular-genetic findings

### 5. Outcomes and their predictive factors

In terms of seizure outcome, 66 patients (90.4%) were completely seizure free and were classified as ILAE 1. No predictive factors were found in this domain (see Supplementary table 2). ASM was withdrawn after surgery in 72.1% (57 children). These patients had a significantly higher preoperative FS-IQ (median 100, range 61 – 124) in comparison to patients who were not ASM-free (median 79.5, range 44 – 106, p = 0.03) (detailed data in Supplementary table 3). Fifty-three patients had neuropsychological evaluation pre- and postoperatively. Eight of them (15.1 %) showed a significant postoperative improvement of FS-IQ, with an increase of more than 10 points after surgery. Only one patient experienced a postoperative decline greater than 10 points. No significant variables predictive of substantial cognitive changes were found. Similarly to the preoperative findings, linear regression analyses showed that younger age at seizure onset (β = +1.21, p = 0.016, R^2^ = 0.108), longer duration of epilepsy (β = –1.84, p = 0.002, R^2^ = 0.19), and a higher number of ASM (β = –5.09, p < 0.001, R^2^ = 0.219) were each significantly associated with lower postoperative FS-IQ (Figures 1D-F). Five patients (6.8%) sustained a permanent postsurgical neurological deficit; two (2.7%) as an unexpected major surgical complication (one patient with hemiparesis due to ischemia of a. basilaris and a second with hemiparesis and aphasia due to ischemia in a. cerebri media) and three (4.1%) as an expected deficit related to lesion localization (hemianopsia due to tumors in occipital region). Ten patients (13.7%) had a minor complication without permanent sequelae. No variables related to unexpected major surgical complications were found. Molecular-genetic findings did not influence outcomes in our cohort.

## IV. DISCUSSION

Our study provides a comprehensive perspective on the effect of epilepsy surgery in a large cohort of children with isolated LEAT and LEAT associated with FCD IIIb. We evaluated seizure outcome, ASM use and withdrawal, postoperative change in FS-IQ and postsurgical complications. Furthermore, we looked for possible predictive factors of the above-mentioned outcomes.

Concerning seizure outcome, our results with 90.4% patients reaching complete postoperative seizure freedom are similar to previously published studies, reporting a seizure-free state in 72 – 90%^18–21^. No factors predicting more favorable seizure outcome were identified in our cohort. Previous studies reported various predictors of favorable postoperative seizure outcome in patients with LEAT, such as shorter epilepsy duration, completeness of resection, unilobar involvement, drug-responsive epilepsy, younger time at surgery, lower frequency of seizures at time of surgery, concordance of EEG findings, and absence of acute postoperative seizures^19,20,22^. Small number of subjects in the non-seizure-free subgroup in our cohort might have led to statistically non-significant results of our analyses.

Our study brings out a unique novel finding, to our knowledge never previously reported in literature on LEAT and epilepsy. Interestingly, we observed a higher proportion of positive results of genetic testing in patients with isolated LEAT, compared to those with LEAT associated with FCD IIIb. Despite a limited cohort size, our findings might possibly suggest a different biological nature of these two groups. While mutation-positive LEAT represent clear neoplasms with tumor-driving pathogenic variants, mutation-negative LEAT associated with FCD IIIb could represent a slightly distinct entity with dysplastic, rather than neoplastic nature^23– 25^. However, these findings need to be confirmed in a larger population and through functional studies. Genetic etiology however does not seem to significantly affect any of the outcome domains, according to our results. Consequently, our findings, in accordance with previous studies, suggest that surgical outcomes in patients with LEAT, isolated or associated with FCD, depend more on surgery- and epilepsy-related variables, primarily the completeness of surgical resection, rather than on the underlying biology of the lesion.

Use of ASM after surgery in patients with LEAT was the subject of investigation in only a few studies. Pellicia et al.^20^ reported 71.6% ASM-free pediatric patients in a cohort of 120 children with LEAT. A smaller proportion of ASM-free patients was reported by Mann et al.^26^, Xie et al.^22^ and Zheng et al.^27^, with 39.3%, 47.3% and 48.6% patients having discontinued ASM. In this context, our results with 78.1% ASM-free patients are very encouraging. To our knowledge, literature on predictive factors of postoperative ASM use is very limited, with only one study reporting a link between focal seizure semiology and ASM withdrawal^28^. Our results do not support this association. In our cohort, a higher probability of ASM withdrawal was related to a higher FS-IQ before surgery. Our findings emphasize the fact that decision for epilepsy surgery should be made as promptly as possible, ideally before the onset of cognitive deterioration, given that epilepsy surgery yields higher chances of post-surgical freedom from seizures and ASM.

In general, neuropsychological outcome is one of the key goals of epilepsy surgery in children. Various authors^29–31^ showed that pre- and postoperative cognition in patients with LEAT depends significantly on its preoperative state, age at seizure onset and duration of epilepsy before surgery. We found similar results in our cohort. A novel finding is the significant inverse correlation between the number of ASM used before surgery and both pre- and postoperative FS- IQ.

Another goal was to identify patients who benefit the most in terms of postoperative cognitive improvement and to detect predictors of this improvement. We have seen a significant cognitive gain in 15.1% of patients but identified no significant variables predicting this improvement. Again, literature is scarce in these data. In a metanalysis by Vasilica et al.^32^ with 911 pediatric patients, there were no significant changes in IQ/DQ after epilepsy surgery in patients with FCD or LEAT.

The risk of surgical complications is a very important issue when we consider epilepsy surgery. In former studies with LEAT, surgical complications occurred in 7.4 – 14.6% of cases^22,33,34^. Previous studies reported a higher age at surgery, multilobar involvement, resections in the rolandic / perirolandic and in insulo-opercular regions as independent risk factors for severe complications. In our cohort, 2.7% had an unexpected major complication and no variables significantly related to them were found. We believe that improvement in presurgical diagnostics, surgical planning and intraoperative monitoring reduced the occurrence of major complications. Finally, there is an increased awareness that timely referral is a crucial predictor of favorable postoperative seizure- and developmental outcomes in epilepsy surgery patients. A clear proof was given by Giulioni et al.^18^, who found an increase of 4% in the risk of recurrence of seizures with each year spent waiting for epilepsy surgery. Moreover, a longer epilepsy duration was associated with neuropsychological deficit in pediatric patients. Therefore, the traditional prerequisite of drug-resistance may be waived in patients with well-defined lesions outside eloquent regions with good anatomo-electro-clinical concordance^34,35^; in fact, more than a quarter of patients in our study were operated on before developing drug resistance.

In summary, the results of our study as well as previous studies and recommendations emphasize the need for an early referral and surgical treatment of children with epilepsy, including caused by LEAT, given that duration of epilepsy and number of ASM used before surgery are the only known predictive factors which can be modified by our therapeutic approach^2^.

## V. CONCLUSION

Epilepsy surgery is highly effective in pediatric patients with LEAT, with the majority achieving freedom from seizures and ASM. Additionally, some patients experience a favorable cognitive outcome after surgery. The incidence of unexpected major surgical complications was very low in our series. Molecular-genetic findings suggest a potentially distinct pathogenesis of isolated LEAT compared to LEAT associated with FCD; however, postsurgical outcomes seem to depend mostly on the possibility of complete removal of the structural epileptogenic lesion.

## Data Availability

All data produced in the present study are available upon reasonable request to the authors

## Acknowledgements

We confirm that we have read the Journal’s position on issues involved in ethical publication and affirm that this report is consistent with those guidelines.

## Glossary

ASM: Antiseizure Medication
DNET: dysembryoplastic neuroepithelial tumor
DRE: Drug Resistant Epilepsy
EEG: Electroencephalography
FBTCS: Focal To Bilateral Tonic-clonic Seizures
FCD Type 1 / 2 / 3: Focal Cortical Dysplasia Type 1 / 2 / 3
FDG-PET: Fluorodeoxyglucose-positron Emission Tomography
GG: ganglioglioma
IEDs: Interictal Epileptiform Discharges
ILAE: International League Against Epilepsy
LEAT: Low-grade epilepsy associated tumors
MOGHE: mild MCD with Oligodendroglial Hyperplasia and epilepsy
MRI: Magnetic Resonance Imaging
SEEG: Stereo Electroencephalography
SISCOM: Subtraction ictal SPECT coregistered to MRI
SPECT: Single Photon Emission Computed Tomography
VEEG: Video Electroencephalography

